# Antibody responses induced by trivalent inactivated influenza vaccine among pregnant and non-pregnant women in Thailand: a matched cohort study

**DOI:** 10.1101/2021.04.07.21255057

**Authors:** Sutthichai Nakphook, Jayanton Patumanond, Manash Shrestha, Kriengkrai Prasert, Malinee Chittaganpitch, Joshua A. Mott, Prabda Praphasiri

## Abstract

**Background:** We compared influenza antibody titers among vaccinated and unvaccinated pregnant and non-pregnant women.

**Methods:** During 1^st^ June – 30^th^ September 2018, four groups of cohort participants - vaccinated pregnant, unvaccinated pregnant, vaccinated non-pregnant, and unvaccinated non-pregnant women were selected by matching age, gestational age, and the week of vaccination. Serum antibody titers against each strain of 2018 Southern Hemisphere inactivated trivalent influenza vaccine (IIV3) were assessed by hemagglutination inhibition (HI) assay on Day 0 (pre-vaccination) and Day 28 (one month post-vaccination) serum samples. Geometric mean titer (GMT), GMT ratio (GMR), seroconversion (defined as ≥4 fold increase in HI titer), and seroprotection (i.e. HI titer ≥1:40) were compared across the study groups using multilevel regression analyses, controlling for previous year vaccination from medical records and baseline antibody levels.

**Results:** A total of 132 participants were enrolled in the study (33 in each of the four study groups). The baseline GMTs were similar for influenza A(H1N1), A(H3N2), and B vaccine strains among all four groups (all p-values >0.05). After one month, both vaccinated groups had significantly higher GMT, GMR, seroconversion, and seroprotection than their unvaccinated controls (all p-values <0.05). The seroconversion rate was over 60% for any strain among the vaccinated groups, with the highest (88.8%) observed against A(H1N1) in the vaccinated pregnant group. Similarly, at least 75% of the vaccinated participants developed seroprotective antibody levels against all three strains; the highest seroprotection was found against A(H3N2) at 92.6% among vaccinated non-pregnant participants. Pregnant women had similar antibody responses (post-vaccination GMT, GMR, seroconversion, and seroprotection) to non-pregnant women for all three strains of IIV3 (all p>0.05).

**Conclusions:** The 2018 seasonal IIV3 was immunogenic against all three vaccine strains and pregnancy did not seem to alter the immune response to IIV3. These findings support the current influenza vaccination recommendations for pregnant women.

## Introduction

Pregnant women are at a higher risk of severe illness from influenza due to physiological and immunological changes and are therefore recommended for influenza vaccination [1-3]. Recent meta-analyses show that influenza vaccination in pregnancy can reduce the incidence of laboratory-confirmed influenza by 53-63%, reduce adverse birth outcomes such as pre-term and low birth weight, and further extend prevention against influenza to their infants [4-6]. The immunogenicity of influenza vaccine is an important measure of vaccine effectiveness, and studies show that concerns related to influenza vaccine effectiveness can be a major barrier to healthcare providers in their recommendations to pregnant women [7]. The data on antibody responses to influenza vaccines, however, remain unclear in pregnancy and are particularly sparse in tropical middle-income countries like Thailand.

Although most studies conducted among pregnant women report adequate humoral immune responses to the trivalent influenza vaccine (IIV3) albeit a lower response to influenza B strain [8-10], only a few studies included a non-pregnant group for comparison [11, 12]. Using vaccinated non-pregnant women as controls, Schlaudecker et al. found that despite similar seroconversion and seroprotection rates, the rise in post-vaccination antibody titers against influenza A(H1N1) and A(H3N2) viruses was diminished in pregnant women [12]. This suggested that immunological changes in pregnancy may modulate the antibody response [12, 13]. In contrast, other observational studies and randomized controlled trials did not report any significant differences in antibody responses to IIV3 between pregnant and non-pregnant women [11, 14].

In Thailand, serious outcomes of influenza including maternal and infant death have been documented [15]. The Thai Ministry of Public Health (MOPH) has recommended seasonal IIV3 for pregnant women since 2009, but the vaccine coverage has been low [16]. Previous studies among Thai pregnant women consistently indicate that their decisions to get vaccinated are mainly influenced by the recommendation of their physicians [17-19], for whom the vaccine efficacy, safety, and immunogenicity data may be important indicators [20]. Extended benefits of maternal vaccination in transferring influenza antibodies to their infants was recently reported by Kittikraisak et al. in a study among Thai pregnant women at a tertiary center in Bangkok [21]. We conducted a prospective cohort study with the main objective of assessing the immune responses induced by IIV3 among both pregnant and non-pregnant women and compared them along with their unvaccinated counterparts.

## Methods

### Study design and setting

This was a prospective, matched cohort study comprising two arms (pregnant and non-pregnant) and four groups – vaccinated pregnant women, unvaccinated pregnant women, vaccinated non-pregnant women, and unvaccinated non-pregnant women in the rural northeastern province of Nakhon Phanom, Thailand (Fig 1). This study was conducted during four months between 1^st^ June – 30^th^ September 2018 as nested research within a larger cohort study assessing influenza vaccine effectiveness among pregnant women in Nakhon Phanom province (Thai Clinical Trials Registry ID: TCTR20201014004). For willing participants, 2018 seasonal southern hemisphere inactivated IIV3 (Influvac®, Abbott Biologicals B.V., The Netherlands) provided free of charge by the Thai MOPH was offered at the provincial hospital (one intramuscular dose of 0.5 ml) containing the following three antigens: A/Michigan/45/2015 A(H1N1)pdm09, A/Singapore/INFIMH-16-0019/2016 A(H3N2), and B/Phuket3073/2013 (Yamataga lineage) [22]. Serum antibody titers against each of the three vaccine strains were assessed on the blood collected from the participants via venipuncture on Day 0 (pre-vaccination) and Day 28 (one month post-vaccination).

**Fig 1.**
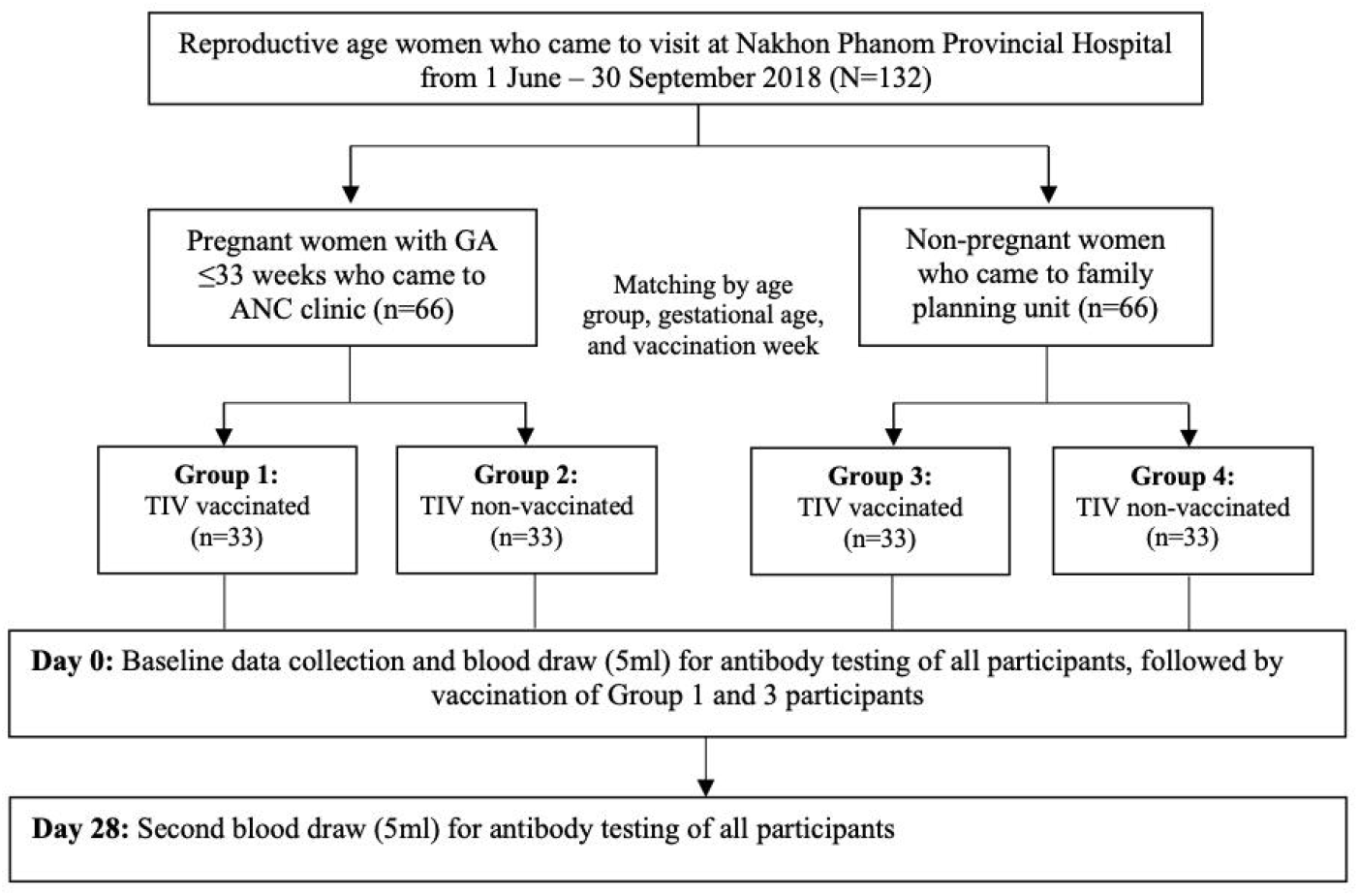
Study flow diagram.

### Participant selection

The study population consisted of women of reproductive age above 18 years visiting Nakhon Phanom Provincial Hospital during the study period. Pregnant women with at least one visit to the hospital’s antenatal care clinic (ANC) were selected if they were Thai nationals, residents of the province since at least June 2018, and had no plans to relocate to another place before giving birth. Pregnant women with gravida more than 33 weeks were excluded to avoid delivery before the second venipuncture, which was scheduled one month after the first venipuncture. Pregnant participants willing and not willing to receive seasonal influenza vaccines were matched by age group, gestational age, and week of vaccination. For comparison, non-pregnant women, both willing and not willing to receive influenza vaccines were selected from the patients visiting the family planning services at the hospital after age-matching with pregnant participants. Their non-pregnant status was confirmed by a negative urine pregnancy test. Any potential participant with an acute illness that could hinder the blood draw procedure was excluded from the study.

### Data collection

After enrollment, demographic data were collected such as age, height and weight (for Body Mass Index (BMI) calculation), gestational age of the baby, number of pregnancies, underlying disease, and smoking status. Histories of the immediate prior year influenza vaccination and presence of any pre-existing medical conditions were obtained from the medical record forms. Particular focus was given to ICD-10 codes for immunosuppressive conditions such as B20 (HIV infection), N18 (Chronic kidney disease), O24 (Gestational DM), E08 (Diabetes mellitus), D89 (Autoimmune disease), C80 (Malignant neoplasm), and C95 (Leukemia). Vaccine strain match data to circulating viruses was obtained from Thai National Institute of Health (NIH) (Supplemental Table 1).

Immediately before vaccination, 5 ml or approximately one teaspoon of blood was drawn from the participants by research nurses using aseptic technique. The blood was collected in a Red Top Vacutainer tube labeled with a unique Study ID Number sticker. The second blood draw was obtained after one month of vaccination. Similarly, for unvaccinated participants, blood samples were drawn on the day of enrollment (Day 0) and after one month (Day 28). For participants who could not travel to the provincial hospital, blood samples were collected by the study nurses at the participant’s home and transported to the provincial hospital on ice packs. Sera were then separated from blood samples using a centrifuge at Nakhon Phanom Hospital Laboratory and stored in a refrigerator at a controlled temperature of 4-8° C for up to 48 hours and then in a freezer at −20° C until used. Serum samples were transported to the Thai NIH laboratory in Nonthaburi every week.

### Serological testing and outcomes

Serum antibody titers were determined by hemagglutination inhibition (HI) assay according to the WHO standard protocol at the Thai NIH laboratory using goose red blood cells as described previously [23, 24]. The laboratory staff were blinded to the participants’ cohort group information. Seroprotection was defined as HI titer ≥1:40 and seroconversion as at least a four-fold increase in antibody titer compared between Day 0 and Day 28 serum samples. The geometric mean titer (GMT) was calculated by taking the antilog of the mean of logarithmically transformed HI titers. Geometric mean titer ratio (GMR) or fold increase was determined as the ratio of GMT of post-vaccination blood by GMT of pre-vaccination blood.

### Sample size calculation

The minimum sample size was calculated based on the Fleiss method [25], using STATA software version 14.2 (StataCorp LP, College Station, TX, USA). At least 28 participants were needed in each of the vaccinated and unvaccinated groups to have 80% power to detect 30% or more seroconversion after vaccination at 5% type I error. To compensate for a possible loss to follow-up, the sample size was inflated by 15% to 33 per group such that the total sample size was 132 (i.e. 33 × 4).

### Statistical analyses

Descriptive statistics were used to enumerate variables such as pregnancy trimester, number of pregnancies, smoking status, and vaccination history. Mean and standard deviation were used to report the central tendency of continuous variables like age, gestational age, and BMI. Estimates of antibody responses (GMT, GMR, seroprotection, and seroconversion) were compared between vaccinated and unvaccinated groups of both pregnant and non-pregnant arms and also between the vaccinated pregnant and non-pregnant women using a multilevel regression, controlling for previous year vaccination history and baseline antibody levels. The use of multilevel regression allowed for adjustment of multiple comparisons by partial pooling and shifting of point estimates and their 95% confidence intervals (CI) [26]. As a sub-analysis, the proportions of vaccinated participants reaching higher HI titers of ≥1:80 and ≥1:160 after one month of vaccination were compared between the pregnant and non-pregnant groups. Statistical analyses were performed using STATA software version 14.2 and the significance was set at p-value <0.05.

### Ethical considerations

The study protocol was reviewed and approved by the Ethical Review Committee of Thammasat University (Ref no. MTU-EC-ES-4-217/60). Approval of local ethics committee of Nakhon Phanom Hospital (No. NP-EC11-No.4/2560) was also received prior to the data collection. Written informed consent was obtained from each participant by research nurses who were not involved in ANC and/or family planning services at the hospital.

## Results

### Participant characteristics

A total of 132 participants (i.e. 66 pregnant and 66 non-pregnant women) were enrolled in the study, with 33 participants in each of the four groups (Fig 1). The baseline demographic characteristics are presented in Table 1. The mean age of the participants was 26.4 years (standard deviation [SD] 5.4 years). The matched frequencies of vaccinated and unvaccinated pregnant participants were 11 (33.3%), 18 (54.6%), and 4 (12.1%) in the first, second, and third trimesters, respectively. Fourteen vaccinated (42.4%) and nine unvaccinated participants (27.3%) were pregnant for the first time. None of the study participants had any pre-existing immunosuppressive medical conditions. BMI and smoking history were comparable across all four groups (p-value >0.05). A similar proportion of vaccinated pregnant and non-pregnant participants had not received influenza vaccination in the previous year (87.9% vs 78.8%, p=0.511) (Table 1).

**Table 1.** Baseline characteristics of study participants (N=132)

| Characteristic | Pregnant women<br>(n=66) |  | p-value | Non-Pregnant women<br>(n=66) |  | p-value | p-value (vaccinated group)* |
| --- | --- | --- | --- | --- | --- | --- | --- |
|  | Vaccination |  |  | Vaccination |  |  |  |
|  | Yes<br>(n=33) | No<br>(n=33) |  | Yes<br>(n=33) | No<br>(n=33) |  |  |
| Age (years) |  |  |  |  |  |  |  |
| Mean (SD)<sup>†</sup> | 26.5 (5.2) | 26.4 (6.0) | 0.931 | 26.8 (5.2) | 26.1 (5.2) | 0.555 | 0.795 |
| Gestational age (in wks.) |  |  |  | - | - | - |  |
| Mean (SD)<sup>†</sup> | 17.4 (7.8) | 19.2 (7.7) | 0.338 |  |  |  |  |
| (Min-Max) | (6-32) | (5-32) |  |  |  |  |  |
| Trimester of pregnancy, n (%)<sup>‡</sup> |  |  |  |  |  |  |  |
| 1<sup>st</sup> trimester (≤13 wks.) | 11 (33.3) | 11 (33.3) | 1.000 | - | - | - |  |
| 2<sup>nd</sup> trimester (14–27 wks.) | 18 (54.6) | 18 (54.6) |  | - | - |  |  |
| 3<sup>rd</sup> trimester (≥ 28 wks.) | 4 (12.1) | 4 (12.1) |  | - | - |  |  |
| Number of pregnancy, n (%)<sup>‡</sup> |  |  |  |  |  |  |  |
| 1 | 14 (42.4) | 9 (27.3) | 0.301 | - | - | - |  |
| ≥2 | 19 (57.6) | 24 (72.7) |  | - | - |  |  |
| BMI |  |  |  |  |  |  |  |
| Mean (SD)<sup>†</sup> | 22.3 (4.6) | 21.1 (2.9) | 0.931 | 22.3 (4.7) | 22.9 (4.3) | 0.545 | 0.795 |
| Smoking history, n (%)<sup>‡</sup> |  |  |  |  |  |  |  |
| Never smoked | 33 (100.0) | 33 (100.0) | 1.000 | 33 (100.0) | 33 (100.0) | 1.000 | 1.000 |
| Influenza vaccination history<sup>‡</sup> |  |  |  |  |  |  |  |
| Received last year (2017) | 4 (12.1) | 1 (3.0) | 0.355 | 7 (21.2) | 2 (6.1) | 0.110 | 0.511 |
| Not received in last year | 29 (87.9) | 32 (97.0) |  | 26 (78.8) | 31 (93.9) |  |  |

Abbreviations: *SD*, Standard deviation; *wks*, weeks; *min*, minimum; *max*, maximum; *BMI*, body mass index.
\**p*-values calculated comparing vaccinated pregnant and vaccinated non-pregnant group
<sup>†</sup>*p*-value calculated using independent *t*-tests, <sup>‡</sup> *p*-value calculated using Exact probability test

### Vaccine strain matching

According to the sentinel surveillance data of the Thai NIH, A/Singapore/ INFIMH-16-0019/2016 A(H3N2) was the dominant strain circulating before the study period and the 2018 seasonal IIV3 vaccine strain matching during the study period for A/Michigan/45/2015 A(H1N1)pdm09, A/Singapore/INFIMH-16-0019/2016 A(H3N2), and B/Phuket/3073/2013 (Yamataga lineage) was 100%, 78.2%, and 100%, respectively (Supplemental table 1).

### Antibody responses to influenza vaccination

The baseline (pre-vaccination) GMTs of the participants were similar for each vaccine strain among all four groups (all p-values >0.05; Table 2). At baseline, the proportion of participants with seroprotective HI titers of ≥1:40 were found to be in the range of 21.2-28.0%, 37.9-48.6%, and 8.9-12.1% for A(H1N1), A(H3N2), and B, respectively.

**Table 2.** Comparison of antibody responses against influenza vaccine strains among the study participants (N=132)

| Antibody response* | Pregnant women<br>(n=66) |  | p-value<sup>†</sup><br>(Pregnancy) | Non-Pregnant women<br>(n=66) |  | p-value<sup>‡</sup><br>(Interaction) |
| --- | --- | --- | --- | --- | --- | --- |
|  | Vaccinated with<br>IIV3 |  |  | Vaccinated with IIV3 |  |  |
|  | Yes<br>(n=33) | No<br>(n=33) |  | Yes<br>(n=33) | No (n=33) |  |
| **Influenza A/Michigan/45/2015**<br>**(H1N1)pdm09** |  |  |  |  |  |  |
| Pre-vaccine GMT (Day 0) | 16.5 | 15.6 | 0.704 | 18.4 | 16.2 | 0.677 |
| Post-vaccine GMT (Day 28) | 171.8 | 14.6 | <0.001 | 106.1 | 17.0 | 0.089 |
| GMR (Fold increase after 28<br>days) | 20.9 | 0.3 | <0.001 | 14.9 | 1.5 | 0.219 |
| Seroconversion (%) | 88.8 | 2.9 | <0.001 | 62.5 | 2.9 | 0.797 |
| Seroprotection (%) (Day 0) | 21.3 | 23.4 | 0.899 | 28.0 | 21.2 | 0.490 |
| Seroprotection (%) (Day 28) | 88.2 | 23.0 | <0.001 | 78.7 | 22.2 | 0.517 |
| **Influenza**<br>**A/Singapore/INFIMH-16-**<br>**0019/2016 (H3N2)** |  |  |  |  |  |  |
| Pre-vaccine GMT (Day 0) | 26.6 | 24.6 | 0.534 | 29.4 | 25.3 | 0.673 |
| Post-vaccine GMT (Day 28) | 195.7 | 26.7 | <0.001 | 241.1 | 27.5 | 0.653 |
| GMR (Fold increase after 28<br>days) | 26.4 | 0.4 | <0.001 | 21.2 | 0.3 | 0.611 |
| Seroconversion (%) | 69.5 | 8.4 | <0.001 | 76.0 | 5.8 | 0.630 |
| Seroprotection (%) (Day 0) | 46.9 | 37.9 | 0.455 | 48.6 | 42.4 | 0.850 |
| Seroprotection (%) (Day 28) | 91.7 | 44.6 | <0.001 | 92.6 | 43.8 | 0.896 |
| **Influenza B/Phuket/3073/2013** |  |  |  |  |  |  |
| Pre-vaccine GMT (Day 0) | 13.6 | 13.2 | 0.677 | 13.3 | 13.3 | 0.758 |
| Post-vaccine GMT (Day 28) | 94.7 | 12.2 | <0.001 | 113.5 | 13.1 | 0.720 |
| GMR (Fold increase after 28<br>days) | 12.3 | <0.1 | <0.001 | 21.1 | 0.5 | 0.133 |
| Seroconversion (%) | 76.9 | 0 | <0.001 | 76.7 | 2.9 | 0.993 |
| Seroprotection (%) (Day 0) | 8.9 | 12.1 | 0.655 | 9.0 | 9.4 | 0.661 |
| Seroprotection (%) (Day 28) | 88.1 | 7.7 | <0.001 | 82.0 | 10.0 | 0.434 |

Abbreviations: IIV3, trivalent inactivated influenza vaccine; GMT, Geometric mean titer; GMR, Geometric mean titer ratio. \*Estimates of antibody response and p-values derived from multilevel regression analyses after controlling for baseline antibody titer and prior influenza vaccination p-value<sup>†</sup> (pregnancy); comparing vaccinated and non-vaccinated participants within pregnant group p-value<sup>‡</sup> (interaction); comparing effect of vaccination across pregnant and non-pregnant participants

After one month, both pregnant and non-pregnant vaccinated women had significantly higher GMT, GMR, seroconversion, and seroprotection in comparison to their unvaccinated control groups (all p-values <0.05; Table 2). Specifically, post-vaccination GMT against A(H1N1), A(H3N2), and B viruses in pregnant women was 171.8 (95% CI 118.8-248.2), 195.7 (95% CI 131.2-291.9), and 94.7 (95% CI 70.4-127.5), respectively; while the same in vaccinated non pregnant women was estimated at 106.1 (95% CI 72.9-154.3), 241.1 (95% CI 161.4-360.0), and 113.5 (95% CI 84.1-153.3), respectively (Fig 2). The GMR was similar among vaccinated pregnant and non-pregnant women for A(H1N1) (20.9 vs 14.9, p=0.219), A(H3N2) (26.4 vs 21.2, p=0.611), and B viruses (12.3 vs 21.1, p=0.133) (Table 2).

**Fig 2.**
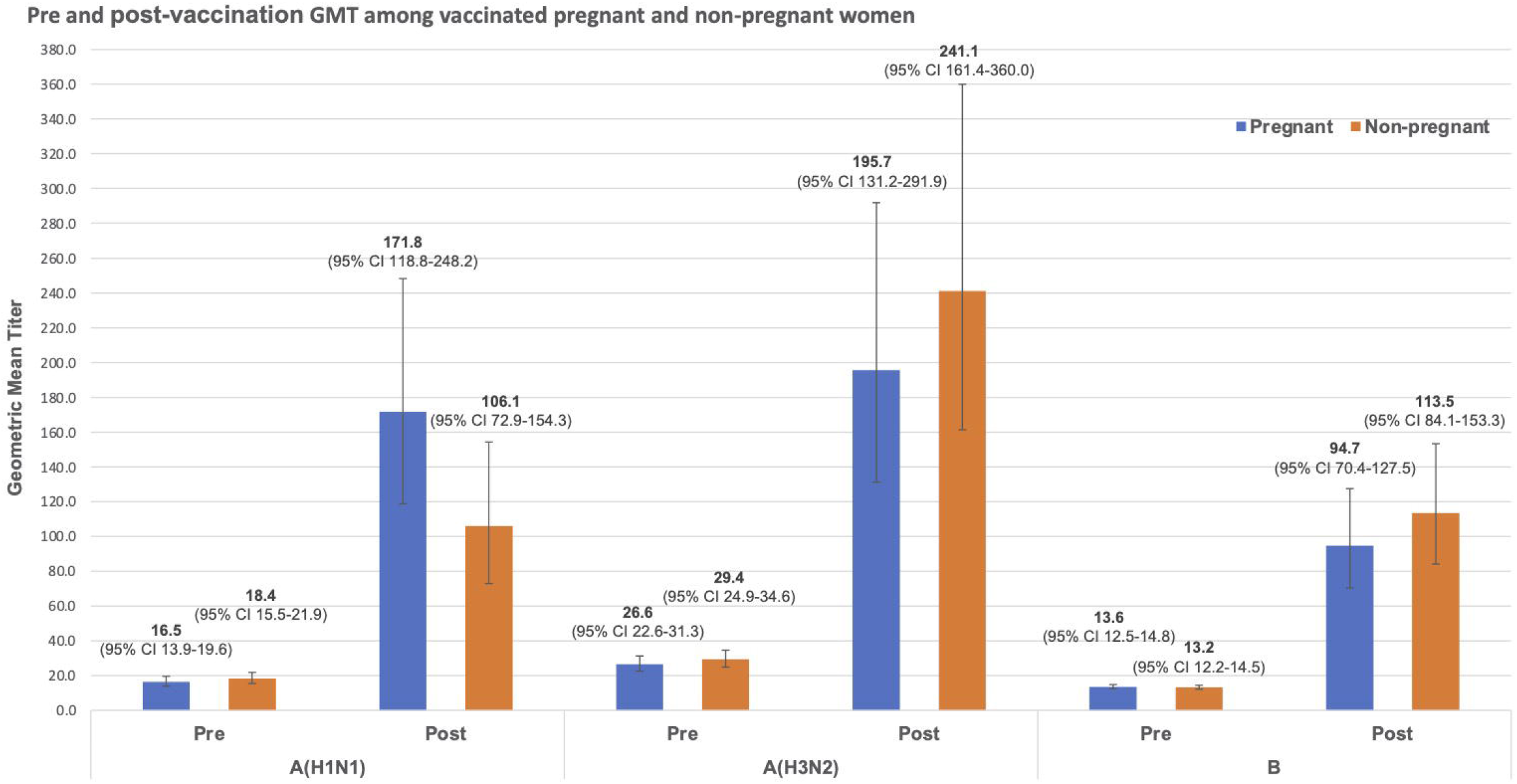
Comparison of influenza antibody geometric mean titers before and after vaccination between pregnant and non-pregnant participants.

The seroconversion rate among vaccinated pregnant women was 88.8%, 69.5%, and 76.9% against A(H1N1), A(H3N2), and B viruses, respectively. The rates were similar among vaccinated non-pregnant women at 62.5%, 76.0%, and 76.7%, respectively (all p-values >0.05; Table 2). At least 78% of the vaccinated participants developed seroprotective antibody levels against all three strains; the highest seroprotection was found against A(H3N2) at 92.6% among vaccinated non-pregnant participants. Similar proportions of both pregnant and non-pregnant participants developed seroprotective HI titers ≥40 after vaccination (p>0.05). When higher cut points of HI titer ≥1:80 and ≥1:160 were used for seroprotection, the proportion of vaccinated pregnant women reaching seroprotective levels for all vaccine strains were in the range of 73-85% and 38-67%, respectively. This was similar among vaccinated non-pregnant women (65-77% and 50-67%, respectively) (Table 3).

**Table 3.** Proportions of vaccinated participants reaching hemagglutination inhibition antibody titers of ≥1:40, ≥1:80, ≥1:160 on one-month post vaccination.

| IIV3 vaccine strain | HI titer cut point | Seroprotection on Day 28 among vaccinated participants* |  | p-value |
| --- | --- | --- | --- | --- |
|  |  | Pregnant (n=33) | Non-pregnant (n=33) |  |
|  |  | % (95% CI) | % (95% CI) |  |
| A/Michigan/45/2015 (H1N1)pdm09 | $\geq 1:40$ | 88.2 (75.0, 101.4) | 78.7 (65.2, 92.2) | 0.517 |
| | $\geq 1:80$ | 73.5 (61.6, 85.4) | 65.9 (53.8, 78.0) | 0.263 |
| | $\geq 1:160$ | 67.7 (55.4, 80.0) | 54.2 (41.7, 66.8) | 0.119 |
| A/Singapore/INFIMH-16-0019/2016 (H3N2) | $\geq 1:40$ | 91.7 (78.7, 104.7) | 92.6 (79.6, 105.7) | 0.896 |
| | $\geq 1:80$ | 84.9 (72.5, 97.2) | 77.6 (65.2, 90.0) | 0.565 |
| | $\geq 1:160$ | 62.7 (50.2, 75.2) | 67.1 (54.6, 79.6) | 0.921 |
| B/Phuket/3073/2013 | $\geq 1:40$ | 88.1 (77.6, 98.6) | 82.0 (71.4, 92.7) | 0.434 |
| | $\geq 1:80$ | 78.7 (67.1, 90.4) | 65.1 (53.3, 76.8) | 0.256 |
| | $\geq 1:160$ | 38.6 (26.7, 50.5) | 50.4 (38.4, 62.4) | 0.466 |

Abbreviations: IIV3, trivalent inactivated influenza vaccine; HI, hemagglutination inhibition; CI, confidence interval.
\*Point estimates, 95% CI, and p-values calculated using multilevel regression, controlling for previous year vaccination and baseline antibody titer.

## Discussion

In our matched cohort analysis, we found that IIV3 induced high humoral immune responses in both pregnant and non-pregnant women compared with their unvaccinated counterparts. In addition, the antibody responses in form of post-vaccination GMT, GMR, and proportions of participants reaching seroconversion and seroprotection levels were not statistically different between the pregnant and non-pregnant women. These findings suggest that pregnancy did not alter immune responses to IIV3 in our study population.

Our results are in stark contrast with that of Schlaudecker et al. who had found significant differences in post-vaccination GMT against influenza A(H1N1)pdm09 and A(H3N2) viruses among pregnant and non-pregnant control group in a similar prospective study [12]. This discordance may be attributed to some differences between the study sample characteristics. The mean age of pregnant women in Schlaudecker et al.’s study was nearly 32 years and almost all of them (97%) had received IIV3 the previous year [12]. Prior vaccination has been associated with lower immune response in subsequent vaccination among pregnant women [27, 28]. In comparison, our participants were younger and likely to be immunologically naïve against the vaccine strains as most of them had not received any influenza vaccines before and therefore could have mounted a higher immune response. Additionally, we controlled for potential confounding factors between the pregnant and non-pregnant women through cohort matching and multilevel regression which may have produced more robust estimates. Similar results of equivalent post-vaccination antibody titers have been found in other studies which have also controlled for baseline differences between pregnant and non-pregnant women [11, 14].

Compared with influenza A(H1N1)pdm09 and B viruses, baseline seroprotection against A(H3N2) was high in our study, conferred possibly by natural infection since it was the dominant circulating strain and prior year vaccination was low. Nonetheless, immune responses against all three influenza strains were strong one month after vaccination in both pregnant and non-pregnant groups, exceeding the European Committee for Medicinal Products for Human Use (CHMP) recommended serological criteria for influenza vaccine for healthy adults aged less than 60 years (i.e. >2.5 GMR, >40% seroconversion rate, >70% seroprotection rate) [29]. In Thailand, the Food and Drug Administration uses the CHMP criteria as a reference for approval of influenza vaccines for public use. However, in 2014, the CHMP adopted a more diversified approach to the measurement and reporting of the immune response to influenza vaccines due to growing concerns of appropriateness of clinical correlation of HI titer ≥1:40 for different subgroups [29, 30]. For example, one study demonstrated that for children, an HI titer >1:110 would be needed for 50% of clinical protection and a titer of 1:330 would be necessary to correlate with 80% clinical protection [31]. This would mean that higher HI titers may be needed among pregnant women to confer clinical protection to their infants, especially up to their first six months during which the newborns are not indicated for influenza vaccination. Since there are no new suggested cut points for pregnant women and an HI titer above ≥1:150 may only correlate with marginal benefits [32], we used HI titers of ≥1:80 and ≥1:160 to further analyze seroprotective levels among our study sample. Our results showed that more than 70% pregnant women reached HI titers ≥1:80 against all three strains and more than 60% reached the titer ≥1:160 after one month of vaccination (except for influenza B), which was not different from those seen among non-pregnant healthy women, denoting a strong immune response.

The vaccine was well-matched with the circulating strains and the antibody responses observed in this study corresponded well with the overall vaccine effectiveness in the larger cohort study which enrolled more than 1,700 participants and estimated the effectiveness of IIV3 against laboratory-confirmed, influenza-associated acute respiratory illness among pregnant women at 65% (95% CI 38%-80%) [33]. These data provide important empirical support to the policy of recommending seasonal influenza vaccination to pregnant women, particularly in countries like Thailand where the vaccine coverage among this group is perennially low. A prior survey revealed that Thai physicians at ANC in public hospitals were more likely to recommend influenza vaccines to pregnant women if they perceived the vaccines to be effective [20]. As healthcare providers’ recommendations are known predictors of IIV3 uptake among pregnant women in Thailand [17, 18], epidemiological evidence of vaccine benefit like effectiveness and immunogenicity in real-world settings may reduce the hesitancy of healthcare providers and aid in their recommendation of IIV3 to pregnant patients.

Despite the strengths of using matched cohorts and controlling for confounders using multilevel regression, there are some limitations in our study. First, the sample size was calculated initially to assess differences between vaccinated and unvaccinated groups which may be small in number and lack sufficient power to draw confirmatory inference between pregnant and non-pregnant women. Consequently, we did not conduct sub-group analyses on the antibody responses by receipt of vaccination in different trimesters of pregnancy. Second, we relied exclusively on HI titers for measuring humoral immunity. Additional measures such as microneutralization assay and induction of plasmablasts may be needed in future studies as they may be more sensitive and specific for pregnant women [11]. Third, the prior vaccination data of the study participants was limited to one year. Although prior vaccinations may affect the immune responses to influenza vaccine, it is less likely that pregnant women have had multiple year vaccinations in Thailand as they are only recommended for seasonal IIV3 in case of pregnancy and the vaccine uptake among pregnant women is less than 1% [16]. Finally, we did not assess the antibody levels in infants which may be an important consideration for the prevention of influenza in infants through maternal vaccination. However, the findings of Kittikraisak et al. suggest that vaccinated Thai pregnant women have higher placental transfer of influenza antibodies to their infants than those unvaccinated [21].

In conclusion, seasonal IIV3 was immunogenic against all three vaccine strains and pregnancy did not seem to alter the immune response to the IIV3. These findings support the current influenza vaccination recommendations for pregnant women. Larger cohort studies with supplementary markers of humoral and cellular immunity may be needed to assess the immune response among pregnant women with subsequent vaccinations.

## Data Availability

All relevant data are within the paper and the supporting files.

## Acknowledgments

We are grateful to the participants and the study nurses at the Nakhon Phanom Provincial Hospital for their consideration and co-operation in this study. We also thank the staff of Thai NIH for their assistance in carrying out the laboratory investigations.

## Data Availability Statement

All relevant data are within the paper and the supporting files.

## Disclaimer

The findings and conclusions in this report are those of the author(s) and do not necessarily represent the official position of the Centers for Disease Control and Prevention or the Agency for Toxic Substances and Disease Registry.

## Author Contributions

Conceptualization: SN KP JP PP

Data curation: SN KP MC

Formal analysis: SN KP JP MS PP

Funding acquisition: KP SN

Investigation: SN KP MC

Methodology: KP JP JAM PP

Project administration: SN KP

Resources: KP

Supervision: JP PP

Validation: MS MC JAM

Visualization: SN MS

Writing – original draft: SN MS

Writing – review & editing: SN JP MS KP MC JAM PP

## Supporting information

**S1 Table. Vaccine strain matching during the study period**

**S2 Dataset and codebook**.

