## Supplemental Table 1 for "Antibody responses induced by trivalent inactivated influenza vaccine among pregnant and non-pregnant women in Thailand: a matched cohort study"

Supplemental Table 1. Vaccine strain matching during the study period*

| Influenza strains | 2018 TIV composition | Circulating strains | Match during the study period |
| --- | --- | --- | --- |
| **A(H1N1)** | A/Michigan/45/2015 (H1N1)pdm09 | A/Michigan/45/2015 (H1N1)pdm09 | 100% |
| **A(H3N2)** | A/Singapore/ INFIMH-16-0019/2016 (H3N2) | A/Singapore/ INFIMH-16-0019/2016 (H3N2)  A/HongKong/4801/2014 (H3N2) | 78.2% |
| **B** | B/Phuket /3073/2013 (Yamataga lineage) | B/Phuket /3073/2013 (Yamataga lineage) | 100% |

**Circulating strains and vaccine strain match data obtained from the sentinel surveillance of Thai National Institute of Health (August 2018 data)*
